# Two Sides of the Spectrum: A Cross-Sectional Study of Autism Diagnosis and Treatment in Kazakhstan from the Perspectives of Parents and Physicians

**DOI:** 10.1101/2025.05.25.25328295

**Authors:** Faye Foster, Akbota Kanderzhanova, Moldir Tazhibay, Akbota Tolegenova, Paolo Colet, Valentina Stolyarova, J Gareth Noble

## Abstract

**Background:** Early diagnosis and intervention for autism spectrum disorder (ASD) are associated with improved outcomes, but access remains uneven in many low- and middle-income countries (LMICs). Kazakhstan has seen increasing awareness of ASD, yet systemic challenges in diagnosis and care persist.

**Objective:** To compare the perspectives of parents and physicians regarding the diagnosis, treatment, and post-diagnostic support of children with autism in Kazakhstan.

**Methods:** A cross-sectional, mixed-methods survey was conducted with 190 parents and 110 physicians across Kazakhstan. Structured questionnaires assessed early symptom recognition, diagnostic experiences, treatment practices, training needs, and beliefs about autism.

**Results:** While 76.6% of parents identified concerns before age three, diagnostic delays were common, and follow-up support was inconsistent. Physicians reported confidence in early identification (86.7%) but low use of standardized tools (26.7%). Notably, 30.0% believed autism could be outgrown with proper treatment, and 36.7% viewed ASD as having a poor prognosis even with early intervention. Nearly half of parents (46.8%) reported being advised to pursue pharmacologic treatment, often in the absence of behavioural therapy. Both groups identified training gaps, limited access to services, and fragmented care coordination as persistent challenges.

**Conclusion:** This dual-perspective study highlights ongoing systemic and perceptual barriers to autism care in Kazakhstan. Addressing misconceptions among clinicians, expanding evidence-based training, and strengthening diagnostic and therapeutic infrastructure are critical for improving outcomes and aligning national practices with global standards.

## Introduction

Autism spectrum disorder (ASD) presents increasing challenges for health systems worldwide, particularly in low- and middle-income countries (LMICs), where diagnostic infrastructure and specialist capacity remain limited (Talantseva et al., 2023; Zeidan et al., 2022). In Kazakhstan, autism services are still developing, shaped by a longstanding tradition of centralized, psychiatry-focused diagnostic practices and limited interdisciplinary training (Mukashova, 2018; Zakirova-Engstrand & Yakubova, 2023). Despite growing awareness, systemic delays in diagnosis and fragmented post-diagnostic support continue to affect families seeking care (An et al., 2020; Yerdessov et al., 2022).

Existing research in Kazakhstan has primarily centred on parental experiences, highlighting barriers such as delayed diagnoses, limited access to behavioural therapies, and inadequate support following diagnosis (An et al., 2020; Kozhageldiyeva et al., 2024). Parents often report high stress levels and dissatisfaction with healthcare interactions, particularly regarding poor communication and lack of guidance after receiving a diagnosis (An et al., 2020; Mukashova, 2018). Financial burdens also remain a major concern, with many families seeking private or alternative therapies due to the unavailability of publicly funded services. Limited access to professional care has also led parents to explore complementary and alternative medicine (CAM), often as a last resort to address their children’s developmental needs (An et al., 2020). This turn toward CAM and private interventions is largely shaped by the stark financial gap between available support and actual care costs: while a standard monthly rehabilitation package may cost around $720 USD, the state-provided disability allowance covers only approximately $76 USD, leaving families to cover the rest out-of-pocket (Pritvorova & Tasbulatova, 2022). As a result, many are forced to forgo essential therapies altogether or rely on non-standard treatment options (An et al., 2020; Pritvorova & Tasbulatova, 2022).

Fewer studies have examined the perspectives of healthcare professionals involved in autism care. Available findings suggest limited use of validated screening tools, uneven knowledge of current diagnostic criteria, and a perceived lack of formal training among physicians (Nukeshtayeva et al., 2022; Somerton et al., 2021). These capacity gaps likely contribute to the diagnostic delays and misalignments reported by caregivers. While physicians and parents are both central to the diagnostic and care process, their experiences and perspectives have not yet been systematically compared in Kazakhstan.

International research indicates that these two groups often interpret delays and care limitations differently (Crane et al., 2018). Parents may attribute delayed diagnoses to physician inaction or misjudgement, while physicians cite diagnostic complexity, insufficient time, and resource constraints (Carbone et al., 2010; Crane et al., 2018). Miscommunication and a lack of mutual understanding can further widen this gap and negatively impact continuity of care (Wiggins et al., 2020).

### Study Aim

This study aims to compare the perspectives of parents and physicians on the diagnosis, treatment, and post-diagnostic support of children with autism in Kazakhstan. By examining experiences related to early symptom recognition, diagnostic timelines, service satisfaction, and perceived barriers to care, the study seeks to identify areas of convergence and divergence between stakeholder groups. The goal is to highlight systemic challenges, such as delays in diagnosis, gaps in provider training, and limited care coordination, and inform future improvements in autism-related healthcare policy and professional education.

## Methods

### Study Design and Approach

This study employed a cross-sectional, mixed-methods survey design to explore and compare the perspectives of two key stakeholder groups involved in autism spectrum disorder (ASD) care in Kazakhstan: parents of children diagnosed with autism and physicians engaged in the diagnosis and management of the condition.

### Survey Instruments

Two structured, researcher-developed questionnaires were designed to reflect the distinct experiences of each respondent group. The development of both instruments was informed by a review of international autism survey tools, relevant literature on diagnostic experiences in low- and middle-income countries, and consultations with clinicians, researchers, and parent advocates in Kazakhstan. Items were adapted to ensure cultural relevance, clarity, and appropriateness for local healthcare contexts and terminology.

- **Parent Survey:** The parent survey comprised 25 multiple-choice questions, one rating scale, and two matrix-format questions. It asked questions relating to parent demographics, early symptom recognition, diagnostic pathways, satisfaction with healthcare encounters, and post-diagnostic support.
- **Physician Survey:** The clinician questionnaire included 52 multiple-choice questions. It asked questions related to demographics, knowledge and beliefs about ASD, diagnostic and treatment practices, perceived barriers to care, and professional training needs.

Both instruments were reviewed by subject matter experts for face and content validity and pilot-tested with a small sample to refine question clarity and relevance.

### Data Collection and Sampling

Data were collected online in two phases: from February to March 2024 for the parent group, and from February to August 2024 for the physician group. Both surveys were offered in Russian and Kazakh.

- **Parent Recruitment:** Participants were recruited using purposive and snowball sampling through rehabilitation centres, social support networks, and digital platforms (e.g., social media, email, and flyers). Recruitment efforts included a personalized video message from a parent of a child with ASD to encourage participation.
- **Physician Recruitment:** A purposive sampling strategy targeted psychiatrists, paediatricians, and paediatric neurologists. Physicians were invited via professional networks, direct emails, and social media outreach.

### Ethical Considerations

Participation in both surveys was voluntary. Informed consent was obtained electronically at the beginning of each survey via a consent confirmation screen. All responses were anonymised, and data were stored securely in accordance with research ethics guidelines. Ethical approval was granted Nazarbayev University School of Medicine (NUSOM) Research Ethics Committee (Re: 2024Feb#04 and 2024Feb#05), Nazarbayev University..

## Data Analysis

Participant responses were analysed using basic descriptive statistics to summarise demographic information, diagnostic timelines, treatment approaches, and satisfaction levels. Comparisons were made between parent and physician groups to explore differences in how early signs of autism were recognised, how long diagnoses took, and what kinds of services were used or recommended. Training participation and interest were also summarised descriptively to capture clinician engagement and unmet needs. For questions that allowed more than one answer, such as reported treatments or barriers to care, response frequencies were calculated based on total selections. This approach captured the range of experiences across both groups. Reported barriers were also ranked by frequency to highlight the most common obstacles to timely and effective care. The analysis focused on identifying where parent and provider perspectives overlapped and where they diverged.

## Results

### Participant Characteristics

Descriptive data on the parent and physician participants are presented in Table 1 and Table 2, respectively. The sample included 190 parents (Table 1), primarily mothers (98%), with an average age of 34 years. Physician participants (Table 2) were predominantly psychiatrists (70.7%) and mostly female (82.7%). Further breakdown of employment status, geographic distribution, and training engagement is detailed in the tables.

**Table 1.** Parent Demographics (n = 190)

| Characteristic | Details |
| --- | --- |
| Total Respondents | 190 (187 parents, 2 grandmothers, 1 guardian) |
| Relationship to Child | 98% mothers (primary caregivers) |
| Gender Distribution | 98% female |
| Age Range | Aged 31–40 years (57.3%) |
| Average Age | Average age: 34 years |
| Employment Status | 57.9% unemployed at time of survey |
| Primary Geographic Location | Astana (46.8%), Almaty (15.2%), Shymkent (7.3%), Karaganda (6.2%), Pavlodar (4.7%), Aktobe, Uralsk, Taldykorgan, Temirtau, and others (combined ~20%) |

**Table 2.** Professional Demographics (n = 110)

| Characteristic | Details |
| --- | --- |
| Total Respondents | 110 (94 Russian-language, 16 Kazakh-language responses) |
| Gender Distribution | 82.7% female |
| Institution Type | 85.2% in state-run institutions (e.g., hospitals and clinics) |
| Professional Specialization | 70.7% psychiatrists, 7.3% paediatricians, 22 % neurologists. |
| Years of Experience | Reported by 82 respondents; approximately 64% had 5–15 years of experience |
| Use of Screening Tools | 26.7% use standardized tools (e.g., M-CHAT), 73.3% rely on ICD-10 or other diagnostic criteria |
| Training Participation | 60% attended lectures, 53.3% participated in conferences, 53.3% self-study |
| Interest in Further ASD Training | 96.7% expressed interest in further ASD-focused training |

### Symptom Recognition and Age at Diagnosis

Parents commonly noticed early signs of developmental concern: 40.8% reported observing symptoms when their child was aged 1–2 years, and 35.8% between 2–3 years. The majority (66.3%) received an official autism diagnosis between the ages of 2 and 5.

Physicians generally expressed confidence in their ability to recognize early signs of autism. A large majority (86.7%) reported feeling comfortable discussing early symptoms with parents, and 73.3% acknowledged that autism can be reliably diagnosed between 18–24 months of age. However, in practice, reliance on clinical criteria such as ICD-10 was high (66.7%), and fewer physicians (26.7%) reported using standardized screening tools like M-CHAT, indicating a gap between theoretical knowledge and applied diagnostic practice.

### Diagnostic Process and Delays

The majority of parents (59.5%) reported initiating the diagnostic process themselves, often driven by concern over symptoms. Other initiators included neurologists (14.6%), relatives (11.7%), and psychiatrists (4.9%). Diagnostic timelines were often prolonged: 24.3% of parents waited 12–24 months from first concern to official diagnosis, while 20.9% waited over two years. Only 28.2% of children received an autism diagnosis at the initial evaluation, while others were referred for additional testing (26.0%), sent to other specialists (16.4%), or reassured that there was no problem (11.3%).

On the physician side, 63.3% reported being authorized to provide an autism diagnosis. However, significant barriers to early detection were identified. Among these, 35.7% cited difficulties recognizing symptoms early in development, and 39.3% highlighted a lack of adequate training. These findings indicate a disconnect between physicians stated diagnostic capacity and parents’ reported experiences of prolonged and fragmented evaluation pathways.

### Post-Diagnostic Support and Resources

Post-diagnosis care varied considerably. While 26.3% of parents reported having a follow-up appointment within one month of diagnosis, and 41.9% within three months, 10.2% had no follow-up at all. Most parents (85.8%) stated that they followed their physician’s treatment recommendations, and 94.8% reported active involvement in creating their child’s care plan.

Despite this engagement, resources provided at diagnosis were often limited. A majority of parents (102) reported receiving only a psychiatric consultation. Sixty-three parents were referred to other specialists, but 35 (out of 177) received no resources whatsoever.

Community support participation was also uneven: only 32.2% of parents were involved in support groups or networks, while 39.5% were not, and 28.2% were unaware such resources existed.

### Treatment Approaches and Interventions

Medication was a common treatment recommendation. Nearly half of parents (46.8%) were advised to start their child on a medication regimen specifically for autism, while 11.5% received medication recommendations only for comorbid conditions. Non-pharmacologic therapies were suggested in 32.1% of cases. 28% of children received no prescription medication.

Among medications, nootropics were most frequently prescribed, reported by 40.4% of parents. Anxiolytics were recommended in 12.5% of cases, and antipsychotics or antibiotics in roughly 3–4%.

Behavioural and developmental interventions were also noted, though less commonly: 19.9% of children received adaptive physical education, and 19.5% participated in Applied Behaviour Analysis (ABA) therapy. These findings reflect a dual emphasis on biomedical and behavioural interventions, although access to comprehensive, non-drug therapies appeared limited.

### Parental Satisfaction and Communication

Parents expressed significant dissatisfaction with multiple aspects of the diagnostic experience. Seventy-two respondents were “very dissatisfied” with how the diagnosis was communicated, and more than half expressed dissatisfaction with the availability of support services (53 “very dissatisfied,” 48 “rather dissatisfied”). In contrast, only 13 parents reported being “very satisfied” with the physician’s knowledge and experience in managing autism.

Overall, dissatisfaction with communication and support resources outweighed satisfaction. This sentiment was mirrored in the widespread call for improved physician education: 94.4% of parents supported the introduction of specialized autism training programmes for healthcare providers.

### Clinical Confidence and Beliefs

Physician beliefs reflected varying levels of confidence and preparedness related to autism identification and care. Most physicians (86.7%) reported feeling confident identifying early signs of ASD, while smaller proportions felt confident explaining the diagnosis to families (56.7%) or making referrals for diagnosis (60.0%). Only 36.7% agreed that existing training was sufficient to support their clinical responsibilities. Notably, 63.3% reported feeling unprepared due to the lack of adapted diagnostic tools in Kazakh and Russian, highlighting a key structural barrier to effective care delivery.

Despite growing awareness of autism, several physician respondents held beliefs that diverge from current evidence-based understanding. Notably, 30.0% of physicians agreed with the statement that children with autism could outgrow the condition with proper treatment, a view not supported by contemporary research. Additionally, 36.7% believed that ASD carries a poor prognosis even when early intervention is provided. These findings highlight outdated beliefs that can affect how diagnoses are made, how families are advised, and what outcomes are expected. Addressing these outdated beliefs through targeted education and updated clinical guidance is essential for improving care pathways and aligning physician perspectives with best practices.

Despite reporting confidence in certain aspects of autism care, many physicians acknowledged gaps in training and preparedness. The desire for further training was nearly unanimous, with 93.3% of physicians agreeing that additional education would improve their ability to care for children with autism. Participation in previous training varied: 60.0% had attended lectures, 53.3% participated in conferences, and 53.3% engaged in self-education. Nearly all respondents (96.7%) expressed interest in further autism-specific training.

### Training Access and Systemic Barriers

Key barriers to accessing such training included time constraints (46.7%) and financial limitations (50.0%). These challenges aligned with parent observations that physicians lacked time and resources. In addition, physicians reported broader system-level obstacles such as limited access to autism specialists (25.0%) and a lack of multidisciplinary support structures (21.4%).

Additionally, none of the physicians reported using the ADI-R, and only 10% reported use of the ADOS-2, tools often regarded as gold standards in international guidelines. Other barriers cited included lack of instrumental support and multidisciplinary team coordination (21.4%), insufficient access to educational resources (17.9%), and time constraints (17.9%). Notably, no respondents indicated lack of support from administration as a limiting factor.

These findings underscore infrastructure and knowledge limitations, rather than organizational opposition, as key barriers to improved autism care.

## Discussion

This study provides the first direct comparison of parent and physician perspectives on autism diagnosis and care in Kazakhstan. While each group interacts with the system from a different vantage point, both identified persistent structural barriers, particularly regarding early diagnosis, treatment access, and post-diagnostic support. These findings reinforce previous single-perspective studies in Kazakhstan and reflect broader patterns observed across other low- and middle-income and post-Soviet settings.

### Early Recognition and Diagnostic Delays

A key area of convergence between groups was the recognition that early identification is possible but inconsistently practiced. Although 76.6% of parents noticed developmental concerns before age three, significant diagnostic delays remained common. Similar to global findings (Carbone et al., 2010; CDC, 2023), parents reported long wait times and a lack of guidance, often initiating the diagnostic process themselves. Physicians, meanwhile, cited difficulty distinguishing early signs and a lack of standardized screening tools—issues previously highlighted by Somerton et al. (2021) and Nukeshtayeva et al. (2022). This mismatch in perceived causes of delay reflects broader research showing how parents may interpret delays as physician reluctance or incompetence, while physicians often attribute them to system-level constraints or diagnostic complexity (Crane et al., 2018).

In Kazakhstan, these delays are likely compounded by legacy diagnostic frameworks and a shortage of trained professionals, especially outside major urban centres (Mukashova, 2018). Although 86.7% of physicians expressed confidence in early recognition, only 26.7% reported using validated tools such as M-CHAT, and 39.3% reported insufficient ASD training. Historically, autism has been classified as a psychiatric disorder only diagnosable by child psychiatrists, which limits diagnostic reach and reinforces urban-rural disparities in access (An et al., 2020; Kosherbayeva et al., 2024).

The persistence of outdated diagnostic criteria, particularly the continued reliance on ICD-10 by 73.3% of physicians, raises concerns as Kazakhstan prepares to transition to ICD-11.

The updated classification system, which came into effect globally in 2022, includes more nuanced definitions of neurodevelopmental disorders that may improve early identification and support (World Health Organization [WHO], 2022). Notably, ICD-11 adopts a lifespan approach and integrates childhood and adolescent disorders into broader diagnostic groupings, enhancing both clinical utility and cultural applicability (First et al., 2019). Effective implementation will require physician retraining, updated national protocols, and clinical guidance to navigate the key changes introduced in ICD-11, including its emphasis on person-centred care and functioning-based assessment models (WHO, 2024).

### Parent–Physician Disconnect

There was a notable mismatch in how parents and physicians understand and navigate the diagnostic process. Parents largely initiated the evaluation themselves (59.5%) and often attributed delays to inadequate physician knowledge or responsiveness. Physicians, in contrast, cited symptom complexity, inconsistent parental concern, and lack of tools as contributing factors, echoing findings in global studies (Crane et al., 2018; Carbone et al., 2010). These disconnects suggest a need for improved communication and shared decision-making practices within ASD care pathways.

### Treatment Gaps and Over-Reliance on Medication

Treatment approaches further revealed gaps between recommended practice and real-world application. Almost half of the parents reported being advised to begin pharmacologic treatment, with nootropics prescribed most frequently, despite limited evidence for their use in ASD. In contrast, evidence-based behavioural therapies such as ABA were accessed by fewer than 20% of children, highlighting issues in service availability and physician training. This pattern aligns with WHO and AAP guidelines, which prioritize psychosocial and behavioural interventions as first-line treatment, particularly in early childhood (Johnson & Myers, 2007; WHO, 2023).

While this study did not directly assess the use of non-evidence-based interventions, the limited mention of behavioural therapies and the reliance on pharmacologic treatments, such as nootropics and antibiotics, raises concerns about alignment with international best practices. Both treatment types are not recommended for core autism symptoms.

Nootropics, though still commonly prescribed in some post-Soviet countries, including Russia and Ukraine, lack robust clinical evidence and are not endorsed by international guidelines. For instance, a double-blind, placebo-controlled trial combining piracetam with risperidone showed some improvement in behavioural symptoms; however, the study’s limitations and the lack of replication studies render the evidence insufficient for widespread clinical endorsement (Tajdari et al., 2008). Similarly, antibiotics have been inappropriately used based on outdated theories linking gut health to autism, despite no proven efficacy.

Recent large-scale studies have found minimal to no association between early-life antibiotic exposure and the development of autism spectrum disorders, suggesting that such treatments should not be employed for managing core autism symptoms (Shao et al., 2024; Wang et al., 2023). These practices are not addressed in Kazakhstan’s current national autism protocol, indicating a need for updated guidelines that align with international standards

### Training Needs and Systemic Barriers

Physician beliefs about autism prognosis also suggest persistent misunderstandings that may influence diagnostic and therapeutic decisions. Thirty percent of respondents believed that children could outgrow autism with proper treatment, while 36.7% believed that ASD carries a poor prognosis even with early intervention. These findings reveal a contradiction: while many physicians expressed confidence and a desire for further training, outdated beliefs about autism prognosis persist. Addressing these misconceptions is essential for ensuring accurate diagnosis, appropriate treatment planning, and effective communication with families. Training initiatives should incorporate not only diagnostic skills but also up-to-date knowledge on autism as a lifelong neurodevelopmental condition and on evidence-based outcome trajectories.

Both parents and physicians emphasized the urgent need for enhanced autism-specific training. Nearly all physicians (96.7%) expressed interest in further education, and 94.4% of parents supported this. These findings echo UNICEF-backed training efforts using tools like ADOS-2 and ADI-R (UNICEF Kazakhstan, 2021), though current reach appears limited. In post-Soviet countries like Kazakhstan, outdated diagnostic models and limited interprofessional collaboration further hinder progress (Zakirova-Engstrand & Yakubova, 2023). Without widespread capacity-building, early diagnosis and intervention goals will remain difficult to achieve.

### Parental Experience and Satisfaction

Many parents reported dissatisfaction with the diagnostic experience, particularly regarding communication and follow-up. Only 26.3% received a follow-up within one month of diagnosis, and 10.2% had no follow-up at all. This lack of structured guidance mirrors findings from other studies in Kazakhstan (An et al., 2020) and underscores a key area for improvement: family-centred care. Better diagnostic communication and referral practices are associated with improved satisfaction and outcomes (Crane et al., 2018; Carbone et al., 2010).

Taken together, our dual-perspective data underscore the need for combined action: implement routine early screening (as recommended by the AAP and CDC), equip front-line physicians with autism-specific training, and build a coordinated care network. International and local bodies agree on these points. WHO and CDC guidance emphasize early identification and intervention (CDC, 2023; WHO, 2023). Local experts also call for expanding multidisciplinary services (An et al., 2020) and embedding autism care in primary health and education systems. Addressing provider capacity, care coordination, and public awareness will be essential for Kazakhstan to move closer to global standards of early diagnosis and sustained family support.

## Conclusion

This study offers the first dual-perspective analysis of autism diagnosis and care in Kazakhstan, highlighting key gaps across early recognition, diagnostic processes, and post-diagnostic support. While parents and physicians agreed on the existence of systemic barriers, their experiences and expectations diverged significantly. Physicians expressed confidence in early identification but reported limited use of validated tools and continued reliance on pharmacologic interventions. Worryingly, many held misconceptions about autism prognosis and the potential for “recovery,” indicating the need to correct persistent belief gaps that could influence treatment decisions and parental guidance. To improve outcomes, Kazakhstan must prioritize evidence-based training, expand behavioural and developmental services, and ensure protocols reflect both best practices and cultural context. Strengthening interprofessional collaboration and aligning physician attitudes with international standards will be essential for building a more inclusive and responsive care system.

## Limitations

This study has several limitations. First, the use of purposive and snowball sampling may limit the generalizability of the findings, particularly to rural regions or underrepresented populations. Second, the reliance on self-reported data introduces the possibility of recall bias or social desirability bias, especially in responses related to physician confidence and training adequacy. Third, while the study captured perspectives from both parents and physicians, the physician sample was predominantly composed of psychiatrists, which may not fully represent the views of other professionals involved in early identification and care, such as paediatricians or primary care providers. Additionally, the study did not include the perspectives of autistic individuals themselves, whose input would be critical in future research to ensure services are responsive to the needs and rights of the autism community.

## Data Availability

The data that support the findings of this study are available from the corresponding author upon reasonable request.

## Author Contributions

**Faye Foster:** Conceptualization, Methodology, Investigation, Writing – Original Draft, Supervision.

**Moldir Tazhibay:** Investigation, Writing – Review & Editing.

**Akbota Kanderzhanova:** Investigation, Writing – Review & Editing.

**Akbota Tolegenova:** Investigation, Writing – Review & Editing.

**Paolo Colet:** Writing – Review & Editing, Supervision.

**Valentina Stolyarova:** Writing – Review & Editing.

**J. Gareth Noble:** Conceptualization, Writing – Review & Editing, Supervision.

